# Empowering Patients with Heart Failure Through Digital Health: The Role of Family Physicians

**DOI:** 10.1101/2025.09.03.25335068

**Authors:** Derya Demirci, Muhammad H. Minhas, Neil G. Barr, Catherine Demers

## Abstract

Heart failure (HF) affects millions worldwide and is associated with high hospital readmission rates. Patients with HF often face challenges such as medication non-adherence, mild cognitive decline, limited healthcare access, and poor knowledge of their condition. Validated digital health tools (DHTs) can help manage HF and prevent symptom worsening. However, it is essential to consider the input of family physicians (FPs) in the development of user-centered DHTs. This study aimed to understand FPs’ attitudes and needs regarding the feasible and effective use of existing self-care DHTs to support older patients with HF. This qualitative study used a user-centered design framework. Semi-structured interviews with twelve FPs were conducted, using persona-case scenarios. The data were analyzed using NVivo 12 software and Braun and Clarke’s thematic analysis method.

The following themes were revealed: *availability of advice in challenging situations, patient clinical data, digital health tool competencies, patient factors, attitudes and preferences toward digital health technology, and the primary care climate*.

This study explores family physicians’ (FPs) needs and expectations in supporting seniors with heart failure (HF) through self-care digital health technologies (DHTs). The findings highlight key challenges FPs face in managing chronic diseases in older adults and emphasize critical considerations for designing HF digital health platforms. Specifically, the study underscores the importance of feasibility and effective integration into primary care, ensuring these platforms align with clinical workflows and enhance patient self-management. These insights can guide the development of user-centered DHTs that support both physicians and patients, ultimately improving HF care in primary care settings.

**Author Summary:** This study aimed to explore family physicians’ (FPs) attitudes and needs regarding the feasibility and effectiveness of supporting older patients with heart failure (HF) using a self-care digital health tool (DHT). The DHT used in this study is CorLibra, a medical device designed to promote HF self-management in the home setting. CorLibra helps patients monitor their weight daily and adjust their diuretic medications accordingly. The tool was specifically developed to address the unique needs of older adults based on extensive qualitative research. Our findings provide valuable insights into how FPs can effectively support patients using such tools to manage chronic conditions like HF.

## Introduction

Heart Failure (HF) is a chronic disease that affects an estimated 750,000 individuals in Canada alone (1). Excess fluid accumulation in the body, which can lead to weight gain, fatigue, swelling in the legs and abdomen, and shortness of breath, are all common symptoms associated with HF (2). Managing HF can be complicated by other chronic diseases, which is the case for one-third of all patients diagnosed with this condition (3). Heart Failure is ubiquitous among the senior population, with more than half of hospitalizations occurring in adults aged 75 years or older (4–7). Unfortunately, the healthcare system faces a significant burden as up to 35% of seniors with HF are readmitted within three months after their initial hospitalization (4–6). The expected healthcare costs associated with HF from 2019/2020 to 2039/2040, there will be 1.69 million heart failure hospitalizations, resulting in a total cost of $19.5 billion for the Canadian healthcare system (8).

Self-care encompasses self-maintenance, self-management, and self-confidence and involves behaviours that maintain physiological stability and response to symptoms when they occur (9). Patient engagement, including self-care, has been shown to improve health outcomes in patients with chronic diseases (10). In patients with HF, self-care includes tasks such as measuring daily weight, symptom monitoring, and adjusting diuretics (11). Systematic reviews found that self-care is effective in preventing hospital readmissions for patients with HF (12–15). In addition, self-care improves patient satisfaction and quality of life (16). However, complex medication regimens, management of comorbidities, health literacy, and frailty can all lead to challenges in self-care (16–19). A model of the HF self-care process can be seen below in **Figure 1** (20).

**Figure 1:**
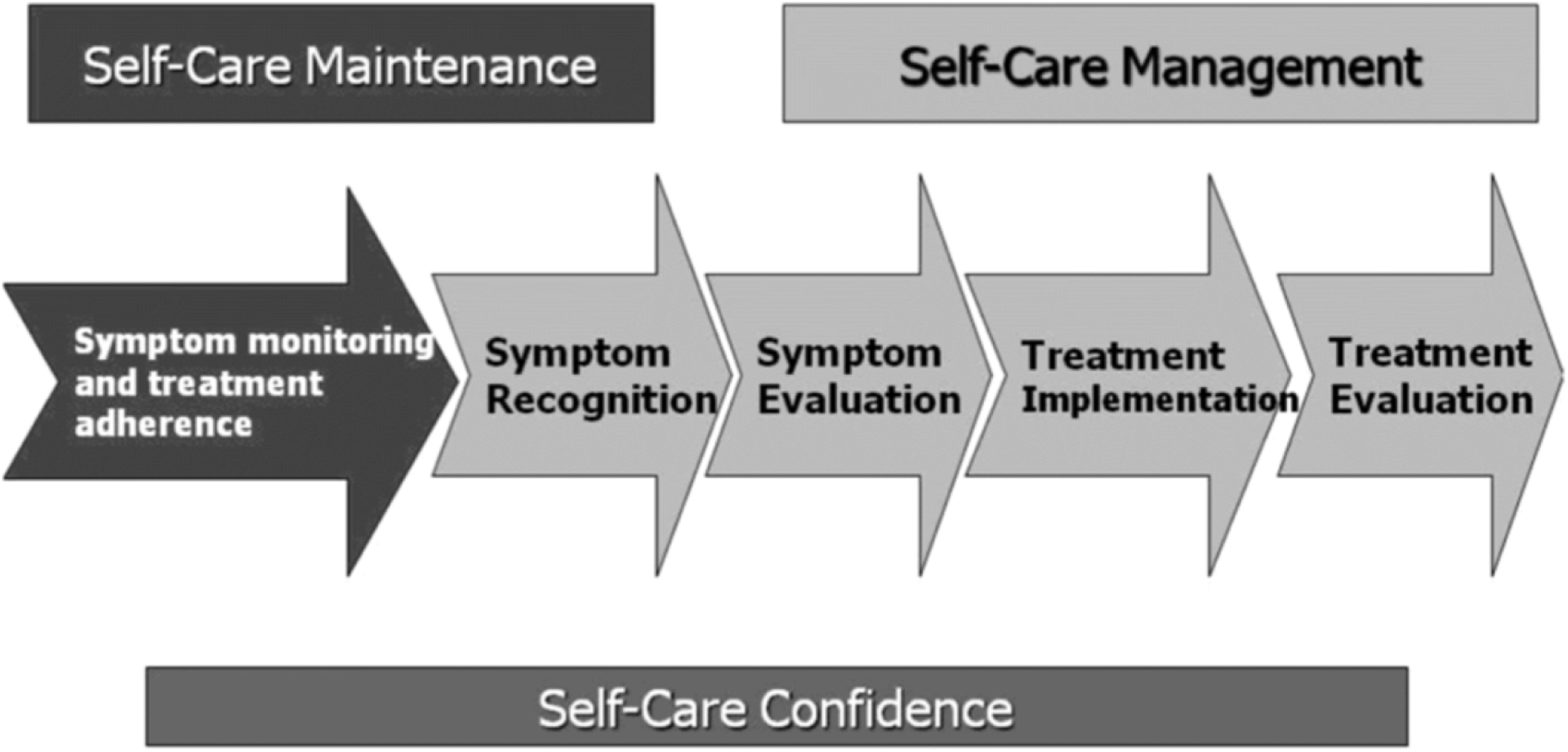
HF Self-Care Process Model.

Currently, team-managed programs catered to patients with HF, such as the Ontario Telemedicine Network’s 6-month virtual care program, have limited ability to support many patients (21). Furthermore, such programs can be costly due to the maintenance of large Personal Health Information Protection Act (PHIPA)-compliant patient databases and the costs of hiring qualified nurse practitioners.

Clinicians, such as family physicians (FPs), often play a critical role in helping patients with HF better understand and manage their health. Patients prefer to see their FPs with whom they have built a strong rapport over specialists (22). However, the attitudes of various stakeholders in managing patients with HF is not well documented (23). A study which included stakeholder attitudes, including FPs, cardiologists, and other healthcare providers (HCPs), found many challenges associated with HF care (23). A major finding was that FPs lacked knowledge and skills associated with HF care (23). Therefore, further efforts are required to identify FPs ’ needs to support and manage patients with HF who utilize DHTs. A literature review found that FPs feel more comfortable managing patients with HF with other healthcare professionals (24). However, barriers such as the lack of sufficient exposure and education needed to manage patients with HF effectively were present (24). Research has also demonstrated the crucial role of diuretics (also known as water pills) in managing HF, as their use can reduce hospitalization rates among patients (25). Yet, this is not a treatment adjustment that all FPs are comfortable making. A study found that FPs felt that cardiologists had unrealistic expectations for managing patients with HF, including making diuretic adjustment (26). Involving FPs in the design of DHTs would also enable them to recommend DHTs to their patients, further promoting self-care (27).

### Study Objective

The objective of this study was to gather family physicians’ (FPs) feedback through persona-case scenario discussions to identify themes and features for integration into a heart failure (HF) self-care tool, CorLibra, to enhance physician support for patients with HF. By understanding FPs’ needs and expectations, the study aims to inform the design of digital health applications that align with clinical workflows and effectively support seniors with HF. Essentially, feedback from FPs might include features that users perceive to be helpful and allow for thorough integration of the tool. Through physician feedback, we can better understand the needs of FPs on how they can better support seniors with HF using CorLibra.

## Materials & Methods

### Study Design

We conducted a qualitative observational study with an experience-based design. An experience-based design is a method which involves a user-focused design process and is often used as a healthcare quality improvement tool (28). It involves patients, caregivers, and stakeholders to identify how healthcare services can be enhanced to provide better care (28). There are four main approaches involving this method which includes: participating action research, user-centered design, learning theory, and narrative-based approaches to change (28). For our context, our participants worked with our staff (study team) to help make changes to improve care for patients with HF, focusing on CorLibra. Participants shared their experiences to inform the development of a more effective solution. This allowed us to maximize patient support and enhance the development of CorLibra. The local Hamilton Integrated Research Ethics Board (HIREB) approved this study (Project #: 14735). Written informed consent was obtained from all participants prior to their inclusion in the study.

### Participants

Participants were male and female FPs between the ages of 24 and 65, who practice in Ontario. Purposeful sampling was completed to get a range of FPs that would allow for information-rich discussions.

### Inclusion Criteria

FPs with a minimum of five years of experience working in primary care were included in the study as it was a reasonable experience level that would ensure that physicians had the experience needed to share thoughtful perspectives. We also ensured that FPs were in non-academic practices as these physicians are not primarily involved in outpatient HF care.

### Exclusion Criteria

FPs with specialties, such as sports medicine or palliative care, were not considered as the study wanted to focus entirely on FPs with the most direct contact with patients without specialty expertise. Participants (FPs) involved in any specialty practices were also not considered, as these family doctors are not primarily involved in outpatient HF care.

## Methods

### Analytic Approach

Braun and Clarke’s inductive thematic analysis approach was used to analyze, identify, reflect and refine emerging themes from the discussion sessions (29). Braun and Clarke’s six-step method requires coder (s) (Derya Demirci, MSc eHealth & Muhammad Hannan Minhas MSc eHealth) to familiarize themselves with the transcripts and then generate initial codes into meaningful groups. Step three involved searching for the actual themes and identifying possible candidates. These candidate themes were reviewed and organized in step four. In step five the themes were defined, named and refined, and lastly there was a report generated for the fully worked-out themes. In addition, Malterud et al’s. (2015) five components, 1) study aim, 2) sample specificity, 3) use of established theory, 4) quality of dialogue, 5) analysis strategy, and guideline on sample size was used as a baseline to determine the sample size for data saturation (30). A qualitative data analysis platform (NVIVO12) was used to systematically analyze the data. To gather feedback on the interview session, an informal feedback questionnaire was developed. The questionnaire aims to capture participants’ thoughts and impressions regarding their experience. Once the reviewers analyzed all of the data, they met collaboratively to review each identified theme, further defining and refining the findings. It was evident that thematic saturation was reached once there were no longer new themes emerging. Moreover, to support each theme, illustrative verbatim quotes were used to allow readers see the evidence underpinning the themes created based on the data analysis.

This study had a relatively narrow objective of using FPs feedback for the further development of Corlibra, and the sample population included participants who had direct primary experience with patients with HF, thus, the sample specificity was also high. Therefore, due to the narrow study aim, dense sample specificity, intense dialogue and case analysis, it was estimated that a large sample size (>30) was not needed to reach data saturation. Furthermore, previous studies indicate that our sample size will provide adequate saturation of coded themes when using structured interviews for qualitative research (29–30). Thus, the projected sample size of twelve participants meets both the student researchers ’capacities (i.e. workload) and data saturation.

We asked FPs to participate in a persona-scenario discussion throughout this study. A persona is an invented person to help represent a type of individual or user. The scenario is then the story or narrative that the persona interacts with (31). Together, these persona scenarios provide insight into FP’s needs and expectations, which helps predict how they would potentially behave and assist patients with HF using self-care tools. The use of persona scenarios also helps individuals identify and connect with what a real person may need (31). It allows them to draw information from another’s experience and connect it to their own. Whether the persona scenario identifies challenges or benefits, it draws different conceptual ideas that may help situations that occur in real life. Discussions were conducted virtually via Zoom and were approximately one hour.

## Results

Interviews were conducted with twelve FPs currently practicing in Ontario. The table below (**Table 1**) outlines FPs sex, years of experience, and practice location (urban vs. rural).

**Table 1:** Characteristics of Study Participants (FPs)

| Characteristics | Participants (FPs)<br>(N=12) |
| --- | --- |
| Sex | • Male=6<br>• Female=6 |
| Experience (Years) | • 5 years or less=3<br>• 6-10 years=7<br>• 10+ years=2 |
| Location of Practice | • Rural areas=3<br>• Urban areas=9 |

Data analysis demonstrated the following major themes: *availability of advice in more challenging situations, patient clinical data, digital health tool competencies, patient factors, attitudes and preferences towards digital health technology, and the primary care climate.* In addition, saturation was reached as there was no further new themes or insights emerging from the data.

### Availability of advice in more challenging situations

Most FPs expressed a general comfort level and positive attitude required to adjust patient diuretics. However, they discussed different aspects of support that would be useful to provide to enable them to access before doing so. Often, a more complex case involving patients with multiple co-morbidities and clinically symptomatic, then FPs are cautious and would prefer to communicate with a specialist.

> **FP:**

> *“If they do not have any renal issues or are a little more straightforward, then I would do the adjustment myself, but if it’s a bit of a nuanced case, then I definitely want some specialist support”*

> **FP:**

> *“I am always worried about pushing a patient into renal failure. I usually I’m just like very conservative with it because I’m scared that I am just going to make things worse”*

Several FPs also expressed the benefits of having access to a cardiologist and having the opportunity to collaborate with them as helpful when providing care for patients with HF.

> **FP:**

> *“I think follow-up with the cardiologist is always best in case they (referring to the patient) need more of a work-up. It depends on how acute the concern is”*

> **FP:**

> *“I have had very helpful cardiologists where I have instructions from them from their consult notes and I can kind of follow that some of the patients actually do know how to make adjustments because they have had very clear instructions”*

Participants also expressed the need for clinical information such as whether the patient is experiencing shortness of breath, their blood pressure, and how much fluid they have been taking before adjusting a patient’s diuretics.

> **FP:**

> *“Does the tool ask questions such as what their blood pressure is? How much fluid they’ve been taking in. Are they short of breath? Do they have chest pain? That needs to be done. I would want to know what the patient actually answered first”*

### Patient Clinical Data

FPs identified a significant advantage to patients utilizing DHT, which is the wealth of data that becomes available to them. In this aspect, FPs favored the HF DHT as it would capture critical information to help manage patients with HF.

> **FP:**

> *“When we are practicing in offices, we see the patients for five-ten minutes, I need to know how they function every day, what their ups and downs are. You cannot treat any human being without you know his lifestyles and this kind of tool will help us do that”*

Participants also expressed the need for clinical information such as whether the patient is experiencing shortness of breath, their blood pressure, and how much fluid they have been taking before adjusting a patient’s diuretics.

> **FP:**

> *“Does the tool ask questions such as what their blood pressure is? How much fluid they’ve been taking in. Are they short of breath? Do they have chest pain? That needs to be done. I would want to know what the patient actually answered first”*

### Digital Health Tool Competencies

FPs identified many factors regarding DHT such as the need to be educated and familiar with the DHTs their patients are using, liability concerns and the importance of having a credible tool supported by evidence-based care. Most FPs expressed a need to know whether their patient is utilizing a HF self-care tool and wanted to be familiar with and educated on how the tool operates. Many of the FPs viewed DHT’s tools from their patients’ attitudes and wanted to be familiar with them to support their patients best. Participants made suggestions that they wanted to be knowledgeable on the DHTs through medical conferences, one-page handouts, links to access more detailed information, or short videos that outline key features. It was also critical for FPs to know their expectations and roles when their patients are utilizing a DHT.

> **FP:**

> *“Knowing first of all that it (referring to the HF self-care tool) exists, how it works on the patient end, how it works on my end, and what the expectations are”*

> **FP:**

> *“I think the doctor needs to be aware of what’s happening just in case you need to kind of check-in”*

Regarding DHTs, FPs expressed a need to have data that highlights the outcomes and benefits such a tool would have for their patients. Providing studies that outline an improvement in patient outcome was deemed critical and also considering who is endorsing the tool.

> **FP:**

> *“But if there was evidence that this tool reduced hospital admissions, mortality, improved quality of life then I think we’re all more motivated to get on board with it because it’s going to change outcomes”*

> **FP:**

> *“I feel, unfortunately, the way our healthcare is right now everybody wants to hear it from the specialist and they take it a lot more seriously than when it comes from a family physician”*

### Patient Factors

Many FPs were supportive of an HF self-care tool if it would help their patients manage their condition. However, they emphasized key factors to remember, such as the patient’s age, access and attitude towards technology, and language barriers. However, these barriers were not significant in introducing and endorsing the tool to their patients as FPs expressed that regardless of patient factors, they would still recommend it because the patients may have a strong support system at home.

> **FP:**

> *“I would recommend such a tool because I never know who’s around in the family and who is their supporting them. But I would certainly consider that there would be frustration and barriers in using them and I would wait for them to let me know what works and what doesn’t work for them”*

> **FP:**

> *“It would have to be something that’s super easy to use because that’s a population that struggles with that kind of thing”*

Participants also expressed features that can be integrated within the tool to ensure that it is adaptable to a diverse patient population. These include multiple language options, simple text and images, regulatory settings for patients with health anxiety and keeping in mind the older population.

> **FP:**

> *“For me, the biggest thing would be a language barrier. So if the tool had lots of pictographs or very simple language that would be helpful”*

> **FP:**

> *“There are some patients that are just like super anxious and they’ll follow their way to a point where it’s unhealthy. For those patients, you’d want to be a little bit careful. Maybe there could be a way to regulate the amount that they’re checking it in those cases”*

### Attitudes & Preferences Towards Digital Health Technology

Participants expressed concern when asked whether they would benefit from having a messaging feature to communicate with patients. Many concerns were raised, especially concerning liability, such as monitoring patients in critical conditions after working hours. Hence, FPs expressed a strong need to have clear guidelines and expectations put into place for patients.

> **FP:**

> *“It becomes a question of what happens if something happens after hours or it’s the weekend and the patient sends you a message and you’re out of the office. I think there would have to be a clear expected time frame”*

> **FP:**

> *“A messaging system has a lot of problems associated with it. First of all, the patients get used to sending messages and they expect an answer back fairly shortly”*

Overall, the majority of FPs had positive attitudes regarding digital health technology and were particularly in favour of tools that would empower their patients.

> **FP:**

> *“You have to embrace technologies that are useful”*

> **FP:**

> *“I’m all about empowering patients with knowledge about their healthcare so that they’re more comfortable and they understand their medical condition”*

In addition, when FPs were asked whether they would want to be informed if their patient is using an HF self-care tool, the majority were strongly in favour.

> **FP:**

> *“If a patient is using a tool, I think it’s important for the doctor to be aware in case a follow-up is needed”*

Specific features that FPs deemed beneficial include items such as providing electronic medical record (EMR) integration, actionable items for patients, sections for FPs to read about what to do in certain scenarios, and access to patients’ answers to symptom questionnaires.

> **FP:**

> *“I would hope that there’s going to be built-in parameters letting us know the alerts we might get and what a standard way to approach it would be”*

> **FP:**

> *“If a patient is using a tool and they’re kind of bringing information to me, it would be nice if the information they’re bringing in is directly actionable”*

> **FP:**

> *“It would be good if there’s educational material that the tool can point to, to be like hey this is what’s going on. Here’s the research behind it. Here’s the data behind it, and here’s who’s using it. It’s been validated this, this and this”*

Overall, FPs strongly emphasized the need for education on managing HF patients utilizing DHTs.

### Primary Care Climate

This was also a key theme seen which referred to the burnout FPs face and the current climate being overwhelmed. Many FPs expressed concerns regarding their workload and that many FPs are leaving the field due to burnout and being overworked. Participants (FPs) expressed the complexity of managing patients with HF and expressed a need to be acknowledged and properly compensated for additional work.

Many FPs expressed concerns regarding their workload and that many FPs are leaving the field due to burnout and being overworked. Participants (FPs) expressed the complexity of managing patients with HF and expressed a need to be acknowledged and properly compensated for additional work.

> **FP:**

> *“I am overwhelmed, if this device has a messaging feature that came to me, I would not be using it. I am so already overwhelmed with patient volume that I just can’t handle it anymore, especially on a topic (referring to heart failure) that I’m not going to have the answer to and I’m going to have to spend time researching or still calling the patient or setting up a visit because I’m not comfortable enough”*

> **FP:**

> *“What would get family physicians to be more active in managing heart failure patients is probably if it was more financially compensated because it’s so complicated”*

## Discussion

Globally, HF impacts millions of individuals (32). It is the number one cause of hospital stay in Canada for people over age 65, accounting for 2 percent of healthcare expenditures (32). The most common symptoms of HF are weight gain, shortness of breath, fatigue and swelling in the legs, ankles and feet (33). Weight gain significantly reflects the worsening of HF. Individuals with HF often struggle to recognize and manage their symptoms. Standard treatment for HF is diuretics, commonly used to relieve symptoms of congestion (34).

Within primary care, FPs are vital in managing patients with chronic diseases such as HF. We must obtain their input to create DHTs to promote self-care for patients with HF. To our knowledge, no study has assessed FPs ’attitudes regarding diuretic adjustments. Our study found that FPs generally all expressed positive attitudes in adjusting patient diuretics for simple cases.

However, with more complex patients, most FPs prefer to consult with a cardiologist or specialist. This also may be because they would like access to advice to be part of the implementation context. Areas for improvement in HF management in primary settings involve adopting collaborative strategies amongst HCPs such as FPs, specialists and nurses (24). Collaboration between FPs and specialists has been found to be more efficient in reducing mortality than primary care alone (24). This emphasizes the importance of establishing strong communication between FPs and specialists to ensure patients receive high-quality care.

DHTs provide important clinical data deemed beneficial by FPs, allowing them to view comprehensive patient data. Early detection of sudden weight gain or weight loss for HF is key to preventing hospital admission and increased mortality (35–36). Regular daily weights are crucial to detect worsening symptoms in patients with HF. Patients using a DHT such as Corlibra will be able to weigh themselves daily, and FPs will be able to view weight trends and important clinical data (37), which was seen positively by FPs. However, obtaining large amounts of data may overwhelm FPs and contribute to poorer care (27). In order to mitigate this challenge, DHTs should clearly outline actionable tasks related to data changes for FPs to reduce complexity and ambiguity (38).

While FPs exhibited favorable attitudes towards DHTs, it is crucial to consider various factors, including HF knowledge, liability, and the need for evidence-based tools and recommendations. The majority of FPs expressed the need to be supported by DHTs to enhance their knowledge, which would allow for better care. There were mixed views about the level of support required. Some expressed thorough support that outlines all the features present within DHTs. Others stated that a general understanding of the DHT would suffice. Many FPs in our study expressed limited knowledge and experience managing HF patients. A study found that providing guidelines or clinical suggestions would help FPs in patient management (39). Supplementary to digital health competencies are liability concerns. Liability concerns are often seen in the majority of DHTs. FPs expressed concerns if patient data or messages were sent during after-hours. This concern can be mitigated by setting time restrictions and training patients on their responsibilities and expectations when using DHTs (40). Patients should be advised to seek medical attention immediately if their symptoms appear to be deteriorating to reduce FPs ’liability concerns. Another possibility is to relay critical patient alerts to a hospital after working hours (40). Ensuring that responsibilities and roles are clearly outlined will help reduce liability concerns expressed by FPs. Lastly, FPs seek DHTs evidence-based supported DHTs that have been shown to improve patient outcomes (40). Therefore, it is crucial for DHTs seeking support from FPs to possess strong scientific backing to ensure its adoption and integration into medical practice (41). While the majority of the FPs viewed DHTs positively if they improved patient outcomes, concerns such as digital literacy, access to technology and patients’ age and cognitive function were still present. Despite the potential challenges, endorsing/recommending a DHT was still seen as feasible by FPs, who believed that patients could rely on their support systems to navigate the process. Nonetheless, it was essential to FPs for DHTs to be simple and catered to diverse population needs. A study which assessed seniors’ perceptions of technology and barriers found that seniors often faced challenges when not provided with guidance/instruction and if the technology was too complex (42). Similar concerns were expressed by FPs in our study hence the emphasis on creating DHTs that are simple, intuitive, and adaptable to seniors.

Health equity was also raised as a concern, with some FPs noting that their patients may face challenges in utilizing DHTs due to insufficient access to technology or language barriers. A solution may be incorporating a validated framework to address health equity concerns. A framework for digital health equity can be incorporated within DHTs (43). According to the framework, there are four key determinants: individual, interpersonal, community and society (37). Individual determinants refer to digital literacy, access to technology and general attitudes toward technology (43). Interpersonal level determinants include implicit technology bias (43). Implicit technology biases refer to the unconscious perceptions that an individual may have towards DHTs, technology and their clinician (43). Disparity populations were seen as less likely to be invited by their clinicians to set up portal accounts due to clinician biases of selecting patients more likely to utilize successfully (43). The FPs in our study demonstrated similar biases in expressing that some patients may succeed more than others when using DHTs. Community-level determinants include community infrastructure, healthy infrastructure, community technology norms and community partners (43). Community infrastructure refers to patients who may not have access to DHTs due to limited internet access. Lastly, societal-level determinants include technology policy, data and design standards (43). These involve federal and local policies to support healthcare technology adoption. Each determinant discussed should be applied to support FPs to ensure they can properly support their patients to utilize DHTs that would improve their conditions. The framework can be seen below in **Figure 2**.

**Figure 2:**
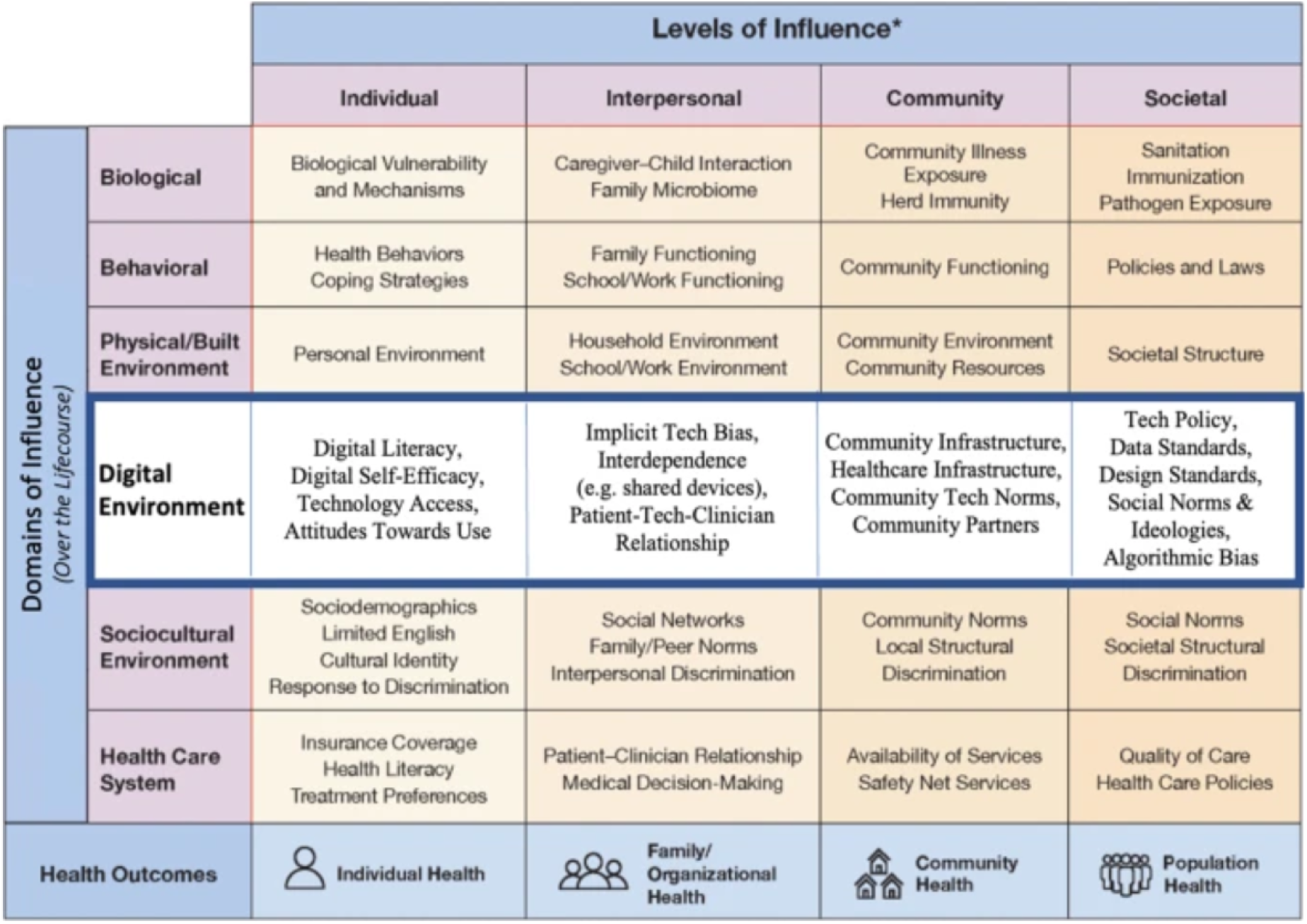
Research Framework for Digital Health Equity.

Lastly, the primary care climate is a significant theme. The family medicine climate in Ontario and across Canada is overburdened and overwhelmed. According to the Ontario Medical Association, 72.9% of physicians stated some level of burnout in 2021 which increased by 66% in 2020 (44). Many of the FPs in our study were cautious about supporting DHTs which could increase their workload. They demonstrated a need to be fairly compensated for their efforts and for the tool to be seamlessly integrated into their workflow was essential such as electronic medical record (EMR) integration. Evidently, due to the current climate, FPs are unable to take on additional responsibilities. Implementing an infrastructure that supports coordinated and integrated primary care is crucial, while also ensuring that the appropriate training, supporting and retention efforts are put into place in human resources (45). Overall, features expressed by FPs in our study highly involved and incorporated educational materials. Some FPs need to gain extensive experience managing HF patients, and limited knowledge may heighten their worries about DHTs. Therefore, DHTs must empower both patients and FPs.

### Limitations

Due to the primary care climate, it was a challenge to recruit a high number of FPs. The original study design was to gain an equal number of FPs who practice in either rural or urban areas. However, due to recruitment limitations this was not possible.

## Conclusion

To effectively promote self-care for patients with chronic conditions like HF, it is essential to consider family FPs ’attitudes and integrate their perspectives into the design and implementation of DHTs. The participants (FPs) highlighted several key factors for successful DHT adoption, including the need for accessible clinical data, ease of use, and consideration of patient-specific factors such as digital literacy. They also emphasized the importance of integrating DHTs into the primary care workflow, which is often constrained by limited resources and high workload. Incorporating FPs’ feedback into the design of DHTs has the potential to improve patient outcomes, enhance medication adherence, and foster greater patient engagement in self-care. By aligning DHTs with the realities of primary care, these tools can better support FPs in managing HF and other chronic conditions, ultimately improving the overall standard of care and reducing healthcare system burdens.

## Data Availability

All relevant data are within the manuscript and its Supporting Information files.

